# Mechanical elements related to the development of patellofemoral pain syndrome, pain intensity, and functional disability: A cross-sectional study

**DOI:** 10.64898/2026.08.26.26361453

**Authors:** Narges Yaghoubi, Mahdi Eghbali, Manijeh Soleimanifar, Fahimeh Hashemirad, Amir Massoud Arab

**Affiliations:** Department of Physical Therapy, Virginia Commonwealth University, Richmond, VA 23298, USA; Department of Physical Therapy, University of Social Welfare and Rehabilitation Sciences, Tehran, Iran; Lifestyle and Health Management Research Group, Academic Center for Education, Culture and Research (ACECR), Tehran, Iran; School of Rehabilitation and Medical Sciences, College of Health Sciences, University of Nizwa, Nizwa, Oman

**Keywords:** Muscle Strength, Patellofemoral Pain syndrome, Posture, Range of Motion

## Abstract

**Background and purpose:** Patellofemoral pain syndrome (PFPS) is a multifaceted condition where proximal, local, and distal factors may contribute to symptoms and limitations. How these factors collectively contribute to PFPS remains poorly understood. Therefore, this study compared proximal, local, and distal mechanical characteristics between individuals with and without PFPS and investigated their association with pain intensity and functional disability.

**Methods:** Eighty participants were included: 40 individuals with unilateral or bilateral PFPS, 40 healthy controls. Isometric muscle strength of hip, trunk, and ankle was assessed using a hand-held dynamometer. Joint alignment (Q-angle, rearfoot angle, pelvic tilt) and muscle flexibility (iliotibial band, hamstrings, quadriceps, gastrocnemius, and soleus) were measured using standard clinical techniques. Pain severity was assessed using a visual analog scale (VAS), and functional disability was evaluated using the Kujala score.

**Results:** Individuals with PFPS showed reduced iliotibial band flexibility, decreased hamstring and soleus length, lower hip abductor strength, and greater anterior and lateral pelvic tilt (all p ≤ 0.02). Multivariate analysis identified reduced iliotibial band flexibility (OR = 7.48) and greater anterior pelvic tilt (OR = 11.75) as independent associates of PFPS. Anterior pelvic tilt predicted pain severity, while anterior trunk muscle strength and Q-angle predicted disability.

**Discussion:** Reduced iliotibial band flexibility and increased anterior pelvic tilt were independently associated with PFPS, while anterior pelvic tilt predicted pain severity and anterior trunk muscle strength and Q-angle predicted functional disability. Clinical assessment and rehabilitation of PFPS should therefore extend beyond the knee to include iliotibial band flexibility, pelvic alignment, and trunk muscle strength.

## 1. Introduction

Patellofemoral pain syndrome (PFPS) is one of the most common clinical entities affecting the lower limbs, particularly the knee joint (Halabchi et al., 2013; Lankhorst et al., 2013; Sisk & Fredericson, 2019; Tkaczyk et al., 2025). This condition is characterized by aching pain in the anterior part of the knee, specifically around or behind the patella (Neal et al., 2019; Sisk & Fredericson, 2019; Smith et al., 2018). Pain is often exacerbated by activities involving flexion of the knee, such as running, squatting, and walking up and down stairs (Lankhorst et al., 2013; Neal et al., 2019; Smith et al., 2018). This syndrome is estimated to affect 22.7% of the general population, with a prevalence that is almost double in women (Neal et al., 2019; Smith et al., 2018). In physically active individuals, particularly runners, 25% of all knee injuries are attributed to PFPS (Halabchi et al., 2013; Smith et al., 2018).

More than half of all patients with PFPS experience poor outcomes five to eight years after treatment, and the condition is now considered a potential precursor to patellofemoral osteoarthritis (Neal et al., 2019, 2025; Petersen et al., 2013; van Leeuwen et al., 2023). The causes of PFPS are complex and involve anatomical, biomechanical, and behavioral factors (Lankhorst et al., 2013; Rathleff et al., 2014; Sisk & Fredericson, 2019).

Although several studies have explored the mechanical factors associated with PFPS, they have focused on specific anatomical regions, such as the knee musculature (Boling et al., 2006; Callaghan & Oldham, 2004; Cowan et al., 2001; Powers et al., 2014), trunk mechanics (Manojlović et al., 2022; Teng & Powers, 2014), and foot posture (Barton et al., 2010).

However, the collective contribution of proximal, local, and distal mechanical factors remains poorly understood, as most studies have examined these variables in isolation rather than simultaneously. Therefore, this study aimed to compare proximal, local, and distal muscle strength, flexibility, and joint alignment characteristics between individuals with PFPS and healthy controls, and to identify which of these factors independently associate with PFPS presence, pain intensity, and functional disability.

## 2. Methods

### 2.1. Study design

This is a cross-sectional observational study compared muscle strength, flexibility, and joint alignment between individuals with PFPS and healthy controls. Within the PFPS group, associations between mechanical variables and both pain intensity and functional disability were also examined. All participants were screened and provided informed consent, approved by the local ethics committee under the supervision of an experienced physiotherapist.

### 2.2. Participants

Eighty participants aged 20-50 years were recruited by convenience sampling from university students and physiotherapy clinic referrals in Tehran and were allocated into PFPS (n = 40) and healthy control (n = 40) groups.

The inclusion criteria were: (1) unilateral or bilateral anterior or retro-patellar knee pain for ≥ 3 months; (2) pain intensity ≥ 3/10 on the VAS; (3) pain during ≥ 3 activities (squatting, jumping, running, kneeling, stair ambulation, prolonged sitting with knee flexed, isometric quadriceps contraction); and (4) pain on patellar compression or palpation tests (e.g., Clarke’s test). The controls had no history of knee pain or lower extremity injury. The exclusion criteria for both groups included other knee pathologies (osteoarthritis or prior surgery), lumbar/hip/ankle pain, ligamentous or meniscal injury, and neurological disorders (Almeida et al., 2015).

### 2.3. Sample size

Pilot data were collected from 16 participants (8 with PFPS, 8 controls). Between-group differences in iliotibial band flexibility showed a large effect size (Cohen’s d > 0.8). Based on these findings, a formal power analysis (two-tailed, α = 0.05, power = 80%) indicated a minimum of 17 participants per group was needed. Given that the study also involved multivariable logistic regression, the sample size was expanded to 40 per group to achieve an acceptable events-per-variable (EPV) ratio of 6.7 for exploratory analysis (Vittinghoff & McCulloch, 2007), while maintaining sufficient power for the between-group comparisons.

### 2.4. Outcome measures

Pain intensity was assessed using the VAS (0-10 cm) (Hawker et al., 2011). Knee function was measured using the Kujala patellofemoral scale (0-100; lower scores indicate greater disability) (Kujala et al., 1993).

Isometric muscle strength of the hip (flexors, extensors, abductors, adductors, internal and external rotators), trunk (anterior muscles, extensors, lateral flexors), and ankle (dorsiflexors, invertors) were assessed using a hand-held dynamometer (HHD) with external stabilization (Martins et al., 2017). The strength values were normalized to body weight.

Joint alignment included the Q-angle and rearfoot angle (standard goniometry) and pelvic tilt in the sagittal and frontal planes (inclinometer).

Muscle flexibility was evaluated for the iliotibial band (Ober test), hamstrings (passive knee extension), quadriceps (Ely’s test), and gastrocnemius and soleus (passive ankle dorsiflexion with the knee extended and flexed, respectively) using standardized clinical tests and goniometer measurements.

### 2.5. Reliability analysis

Intra-rater reliability was assessed in a pilot sample of 16 participants. All outcome measures demonstrated excellent reliability (ICC > 0.75; table 1).

**Table 1.**
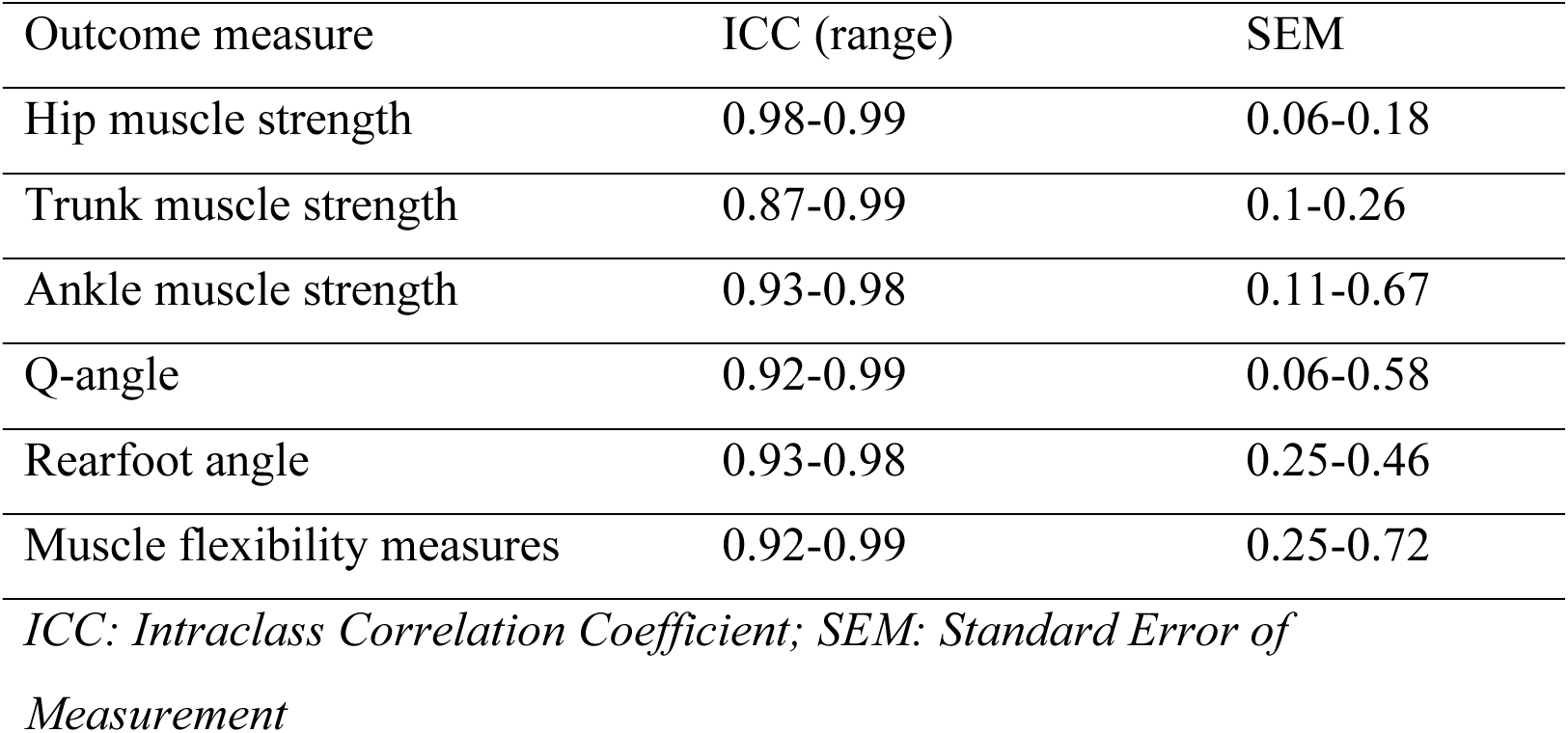
Reliability table.

| Outcome measure | ICC (range) | SEM |
| --- | --- | --- |
| Hip muscle strength | 0.98-0.99 | 0.06-0.18 |
| Trunk muscle strength | 0.87-0.99 | 0.1-0.26 |
| Ankle muscle strength | 0.93-0.98 | 0.11-0.67 |
| Q-angle | 0.92-0.99 | 0.06-0.58 |
| Rearfoot angle | 0.93-0.98 | 0.25-0.46 |
| Muscle flexibility measures | 0.92-0.99 | 0.25-0.72 |

*ICC: Intraclass Correlation Coefficient; SEM: Standard Error of Measurement*

### 2.6. Procedures

All measurements were performed by a single experienced physiotherapist in a standardized laboratory setting, with three trials recorded and averaged for each measurement.

#### 2.6.1. Muscle strength assessment

Isometric strength was measured using a belt-stabilized HHD (North Coast Medical Inc., Morgan Hill, CA, USA) with external straps (Ireland et al., 2003; Martins et al., 2017) following standardized manual muscle testing protocols (Kendall et al., 1993). Each contraction was a maximum voluntary isometric contraction maintained for five seconds. Hip muscle groups (flexors, extensors, adductors, abductors, and internal and external rotators) were tested in standardized positions according to established protocols (Cichanowski et al., 2007; Ireland et al., 2003; Magalhães et al., 2010; Nakagawa et al., 2012). The ankle invertors and dorsiflexors were assessed in the supine position (de Moura Campos Carvalho E Silva et al., 2014). Trunk anterior muscles, extensors, and lateral flexors were assessed using handheld dynamometer-based protocols with appropriate stabilization (Nakagawa et al., 2015) (Figure 1).

#### 2.6.2. Joint alignment measurements

The Q-angle was measured in the supine position with the knee at approximately 10° flexion using a universal goniometer (fulcrum over the patella, arms aligned with the ASIS and tibial tuberosity) (Almeida et al., 2016). The rearfoot angle was assessed in the prone position with the subtalar joint in neutral position using a goniometer aligned with the lower leg and calcaneal midlines (Levinger & Gilleard, 2004; Powers et al., 1995). Pelvic tilt was assessed in the standing position using a pelvic inclinometer (Hagins et al., 1998; Herrington, 2011): one pointer over the ASIS and the other over the PSIS for anterior–posterior tilt, and both pointers over the ASIS landmarks for lateral tilt. All measurements were performed in triplicate and averaged.

#### 2.6.3. Flexibility and muscle length assessment

Iliotibial band flexibility was assessed using the Ober test with an inclinometer at the lower iliotibial band (Ober & Peltier, 1935; Reese & Bandy, 2003). Hamstring length was assessed using passive knee extension in the supine position at 90° hip flexion, with a goniometer over the lateral femoral condyle (Hopper et al., 2005; White et al., 2009; Youdas et al., 2005). Quadriceps length was assessed using Ely’s test (passive prone knee flexion with pelvic stabilization); the end range was the point of firm resistance or the onset of pelvic movement (Magee, 2021). The gastrocnemius and soleus lengths were assessed using passive ankle dorsiflexion in the supine position with the knee extended and at 90° flexion, respectively, and a goniometer was positioned over the lateral malleolus (Kendall et al., 1993; Magee, 2021).

### 2.7. Statistical analysis

IBM SPSS Statistics v31.0 was used for all the analyses. Descriptive data are reported as mean ± SD. The Shapiro–Wilk test was used to check data normality. For between-group comparisons, independent t-tests were applied to continuous variables and chi-square tests to categorical variables. Variance homogeneity was assessed using Levene’s test.

Variables significantly different between groups were entered into a multivariable binary logistic regression to identify independent mechanical factors associated with PFPS, and multicollinearity was assessed prior to model entry. Odds ratios (ORs) and 95% confidence intervals (CIs) were calculated, and the number of predictors was limited to maintain an adequate event-to-variable ratio.

Within the PFPS group, associations between mechanical variables and VAS and Kujala scores were examined using Spearman’s correlation analysis, and significant variables were entered into linear regression models. A stepwise approach was used for the disability model to prevent overfitting. Statistical significance was set at P < 0.05.

## 3. Results

### 3.1. Participants

Eighty participants were included in the study (40 with PFPS and 40 controls). In the PFPS group, 11 patients had unilateral symptoms and 29 had bilateral symptoms; the more symptomatic limb was used for bilateral cases. The control limb values were averaged. The groups were well matched for demographic variables (Table 2).

**Table 2.**
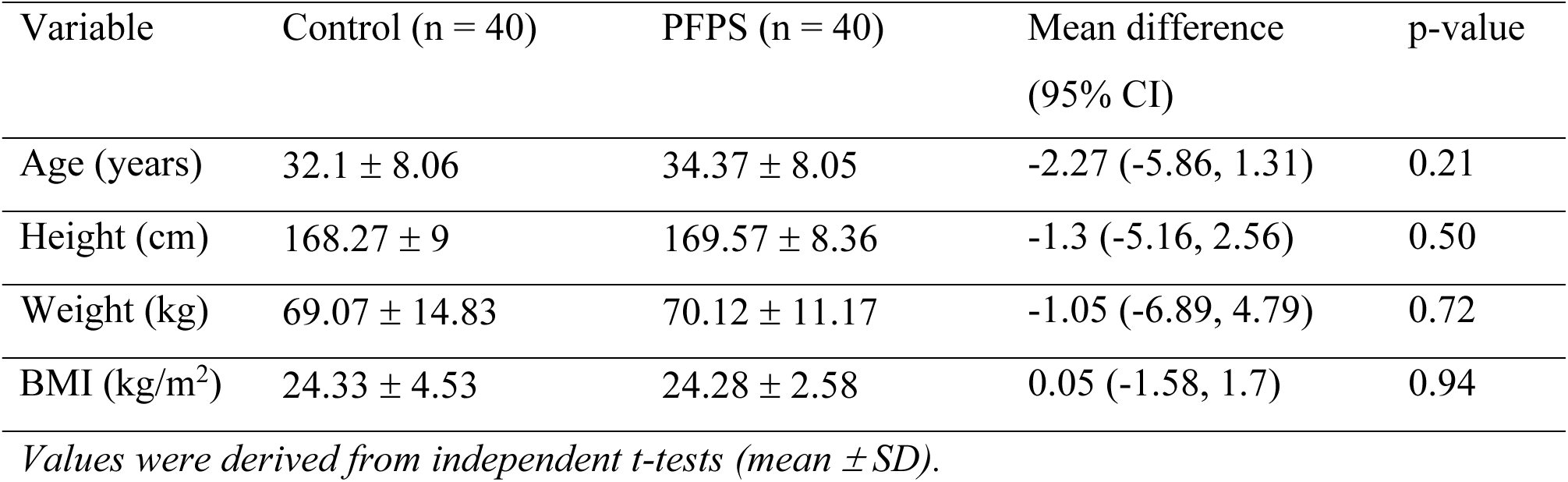
Participants characteristics of the PFPS and control groups.

| Variable | Control (n = 40) | PFPS (n = 40) | Mean difference<br>(95% CI) | p-value |
| --- | --- | --- | --- | --- |
| Age (years) | 32.1 ± 8.06 | 34.37 ± 8.05 | -2.27 (-5.86, 1.31) | 0.21 |
| Height (cm) | 168.27 ± 9 | 169.57 ± 8.36 | -1.3 (-5.16, 2.56) | 0.50 |
| Weight (kg) | 69.07 ± 14.83 | 70.12 ± 11.17 | -1.05 (-6.89, 4.79) | 0.72 |
| BMI (kg/m<sup>2</sup>) | 24.33 ± 4.53 | 24.28 ± 2.58 | 0.05 (-1.58, 1.7) | 0.94 |

*Values were derived from independent t-tests (mean ± SD).*

### 3.2. Pain and disability

Individuals with PFPS reported significantly greater pain intensity (d = 3.37) and lower Kujala scores (d = 1.63) than those in the control group (Table 3).

**Table 3.** Pain and disability comparison between groups.

| Variable | Control (n = 40) | PFPS (n = 40) | Mean Difference (95% CI) | Effect Size (Cohen's $d$) | p-value |
| --- | --- | --- | --- | --- | --- |
| VAS (0-10) | 0.0 | $4.42 \pm 1.85$ | 4.42 (3.83, 5.01) | 3.37 | < **0.001** |
| Kujala (0-100) | $98.57 \pm 3.94$ | $83.1 \pm 12.82$ | 15.47 (11.20, 19.74) | 1.63 | < **0.001** |

*Values are mean ± SD; derived from independent t-tests.*

### 3.3. Between-group comparisons of mechanical variables

Significant differences were observed in hip abductor strength, iliotibial band flexibility, hamstring and soleus lengths, and pelvic alignment (Table 4). No significant differences were found in other strength measures, Q-angle, rearfoot angle, or quadriceps/gastrocnemius flexibility (p > 0.05).

**Table 4.** Between-group comparisons of mechanical variables.

| Variable | Control | PFPS | Mean Difference (95% CI) | Cohen's $d$ | p-value |
| --- | --- | --- | --- | --- | --- |
| Hip flexor strength | $13.9 \pm 5.89$ | $13.19 \pm 6.4$ | 0.71 (-2.02, 3.45) | 0.11 | 0.60 |
| Hip extensor strength | $9.48 \pm 4.08$ | $8.18 \pm 3.45$ | 1.3 (-0.37, 2.98) | 0.34 | 0.12 |
| Hip adductor strength | $8.82 \pm 4.82$ | $7.27 \pm 2.87$ | 1.54 (-0.22, 3.32) | 0.39 | 0.08 |
| Hip abductor strength | $11.42 \pm 6.45$ | $8.85 \pm 2.87$ | 2.56 (0.32, 4.8) | 0.51 | **0.02\*** |
| Hip internal rotator strength | $7.31 \pm 2.4$ | $6.66 \pm 1.96$ | 0.65 (-0.32, 1.63) | 0.29 | 0.18 |
| Hip external rotator strength | $7.01 \pm 3.04$ | $6.73 \pm 2.09$ | 0.28 (-0.88, 1.44) | 0.1 | 0.62 |
| Ankle dorsiflexor strength | $5.61 \pm 3.46$ | $5.14 \pm 2.41$ | 0.47 (-0.85, 1.8) | 0.15 | 0.48 |
| Ankle invertor strength | $3.92 \pm 2.05$ | $3.83 \pm 1.73$ | 0.08 (-0.75, 0.93) | 0.04 | 0.83 |
| Anterior trunk muscles strength | $4.84 \pm 2.18$ | $4.56 \pm 1.97$ | 0.28 (-0.64, 1.2) | 0.13 | 0.54 |
| Trunk extensor strength | 4.19 ± 2.51 | 4.72 ± 2.17 | -0.52 (-1.57, 0.51) | -0.22 | 0.31 |
| Trunk lateral flexor strength | 8.51 ± 3.32 | 8.75 ± 2.88 | -0.24 (-1.63, 1.13) | -0.07 | 0.72 |
| Q-angle | 14.16 ± 2.39 | 13.63 ± 2.09 | 0.52 (-0.47, 1.52) | 0.23 | 0.29 |
| Iliotibial band flexibility | -20.61 ± 3.92 | -14.78 ± 2.93 | -5.83 (-7.37, -4.29) | -1.68 | **< 0.001\*** |
| Quadriceps length | 140.02 ± 4.8 | 138.15 ± 3.98 | 1.87 (-0.08, 3.84) | 0.42 | 0.06 |
| Hamstring length | -24.15 ± 8.87 | -29.55 ± 5.50 | 5.39 (2.09, 8.68) | 0.73 | **0.002\*** |
| Gastrocnemius length | 92.37 ± 4.07 | 91.75 ± 4.08 | 0.62 (-1.18, 2.44) | 0.15 | 0.49 |
| Soleus length | 107.18 ± 4.08 | 104.12 ± 4.13 | 3.05 (1.22, 4.88) | 0.74 | **0.001\*** |
| Rearfoot Angle | 11.47 ± 2.07 | 11.28 ± 2.09 | 0.18 (-0.74, 1.11) | 0.09 | 0.68 |
| Anterior pelvic tilt | 2.92 ± 0.97 | 6.15 ± 1.45 | -3.22 (-3.77, -2.67) | -2.59 | **< 0.001\*** |
| Lateral pelvic tilt | 2.5 ± 1.28 | 3.72 ± 1.75 | -1.22 (-1.9, -0.54) | -0.79 | **< 0.001\*** |

Values are mean ± SD. \* $p < 0.05$ . Strength values were normalized to body weight.

### 3.4. Logistic Regression

The multivariable binary logistic regression model was statistically significant (χ²(6) = 95.05, p < 0.001), with good calibration (Hosmer–Lemeshow p = 1.00), strong discrimination (Nagelkerke R² = 0.93), and 95% sensitivity and specificity at a 0.50 cutoff. Given the cross-sectional design and sample size, these estimates should be interpreted cautiously.

Reduced iliotibial band flexibility (OR = 7.48, 95% CI = 1.48–37.71, p = 0.015) and greater anterior pelvic tilt (OR = 11.75, 95% CI = 2.10–65.78, p = 0.005) were independently associated with PFPS. Hip abductor strength, hamstring length, soleus length, and lateral pelvic tilt were not independently associated (Table 5).

**Table 5.** Multivariable logistic regression-mechanical factors associated with PFPS.

| Predictor | OR (Exp(B)) | 95% CI | p-value |
| --- | --- | --- | --- |
| Hip abductor Strength | 0.92 | 0.50 – 1.69 | 0.779 |
| Iliotibial band flexibility | **7.48** | 1.48 – 37.71 | **0.015** |
| Hamstring Length | 0.98 | 0.78 – 1.24 | 0.868 |
| Soleus Length | 0.85 | 0.57 – 1.29 | 0.449 |
| Anterior pelvic tilt | **11.75** | 2.10 – 65.78 | **0.005** |
| Lateral pelvic tilt | 1.30 | 0.46 – 3.67 | 0.617 |

*\*p < 0.05. An OR > 1 indicates increased odds of PFPS.*

### 3.5. Association with pain and functional disability

In the PFPS group, Spearman’s correlations showed that pain severity (VAS) was positively correlated with anterior pelvic tilt (ρ = 0.45, p = 0.003) and negatively correlated with hip extensor strength (ρ = −0.36, p = 0.025). Kujala scores were negatively correlated with anterior pelvic tilt (ρ = −0.44, p = 0.005) and lateral pelvic tilt (ρ = −0.35, p = 0.029) and positively correlated with Q-angle (ρ = 0.32, p = 0.045), hip internal rotator strength (ρ = 0.32, p = 0.044), ankle dorsiflexor strength (ρ = 0.36, p = 0.024), ankle inverter strength (ρ = 0.35, p = 0.029), and anterior trunk muscle strength (ρ = 0.42, p = 0.007).

Multiple linear regression confirmed anterior pelvic tilt as the sole independent predictor of pain severity (β = .460, p = .005; R² = .283, adj. R² = .244; F(2,37) = 7.30, p = .002). Anterior trunk muscle strength (β = .521, p < .001) and Q-angle (β = .327, p = .019) were independent predictors of functional disability (R² = .345, adj. R² = .309; F(2,37) = 9.73, p < .001).

## 4. Discussion

This study set out to determine which proximal, local, and distal mechanical factors are associated with PFPS presence, and whether these factors also predict pain intensity and functional disability. The main findings were that iliotibial band tightness and anterior pelvic tilt independently distinguished individuals with PFPS from healthy controls. Within the PFPS group, anterior pelvic tilt was the strongest predictor of pain intensity, while anterior trunk muscle strength and Q-angle accounted for functional disability.

### Iliotibial band flexibility

Among the variables examined, reduced iliotibial band flexibility showed the strongest association with PFPS presence, with an odds ratio of 7.48. This is in line with earlier work reporting altered lateral soft-tissue mechanics in people with anterior knee pain (Hudson & Darthuy, 2009; Puniello, 1993; Winslow & Yoder, 1995). Mechanically, a tight iliotibial band may pull the patella laterally, increasing lateral tracking forces and compressive stress within the patellofemoral joint — an effect that would be particularly pronounced during knee flexion (Merican & Amis, 2009; Waryasz & McDermott, 2008). Clinically, this may reflect chronic adaptive changes in the lateral thigh tissues, underscoring the importance of addressing the contributions of the lateral kinetic chain in PFPS management (Puniello, 1993; Sanchis-Alfonso, 2014; Sisk & Fredericson, 2019).

### Anterior pelvic tilt

In the present study, greater anterior pelvic tilt emerged as an independent associate of PFPS, pointing to a proximal mechanical contribution to patellofemoral joint loading. This finding is noteworthy because anterior pelvic tilt does not act directly on the patella. Rather, it likely reflects a broader lumbopelvic postural or neuromuscular pattern that modifies femoral kinematics and joint stress during weight-bearing activities (Crossley et al., 2018). This interpretation is consistent with evidence suggesting that altered pelvic orientation affects load distribution throughout the lower extremity (Petersen et al., 2013; Sanchis-Alfonso, 2014), and aligns with the broader clinical shift toward assessing proximal factors in PFPS management (Piva et al., 2005; Powers, 2010).

### Strength and flexibility

Although hip abductor weakness and reduced hamstring and soleus lengths were present in the PFPS group, these variables were not independently associated with PFPS in the multivariate model. Hip abductor weakness is widely reported in patients with PFPS (Neal et al., 2019; Rathleff et al., 2014), and hip strengthening is emphasized in rehabilitation (Barton et al., 2015; Lack et al., 2015). These findings suggest that such impairments may be secondary or mediated by other mechanical factors.

### Joint alignment

No significant between-group differences were observed in the Q-angle or rearfoot angle, consistent with systematic reviews questioning the clinical relevance of static alignment measures in PFPS (Lankhorst et al., 2013; Neal et al., 2019). Static alignment may not capture the dynamic loading conditions relevant to patellofemoral pain (Petersen et al., 2013; Powers, 2010).

### Pain and disability

Anterior pelvic tilt was most strongly associated with pain severity, whereas anterior trunk muscle strength and Q-angle were independent predictors of functional disability.

## 5. Implications for Physiotherapy Practice

The results of this study have direct implications for clinical practice. Since iliotibial band flexibility and anterior pelvic tilt were independently associated with PFPS, these factors should be part of routine clinical assessment. The Ober test and a standard inclinometer are sufficient for this purpose and are available in most clinical settings. Because anterior pelvic tilt also predicted pain severity, addressing pelvic alignment should be included in the treatment plan from early stages. Additionally, to improve functional outcomes, rehabilitation should incorporate exercises targeting anterior trunk muscle strength and strategies to reduce Q-angle.

## 6. Limitations

Several limitations should be acknowledged. The cross-sectional nature of the study means that causal relationships cannot be established, and we do not know whether the mechanical characteristics identified in this study developed before or after PFPS development. Second, all measurements were taken under static or quasi-static conditions; therefore, it is unclear whether or how movement patterns that occur during dynamic functional tasks are accounted for in PFPS. Third, clinical methods were used instead of 3D gait analysis using instrumented equipment, which may provide less detail in terms of movement biomechanics. Although we used standardized methods to collect data and reduce potential bias, clinical goniometry and manual muscle testing are also subject to examiner biases. Further validation of the current findings using a larger, independent sample is required.

## 7. Conclusion

Restricted iliotibial band flexibility and increased anterior pelvic tilt were independently associated with PFPS presence. Anterior pelvic tilt was the only factor that predicted the pain severity. Anterior trunk muscle strength and Q-angle were the only factors that predicted functional disability in this study. These findings highlight the importance of a comprehensive assessment of various mechanical factors at the proximal, local, and distal levels in patients with PFPS. Further studies are required to assess the longitudinal relationships between these factors.

## Acknowledgments

We are grateful to all the participants for generously contributing their time and experiences, which were essential for the success of this study.

## Data Availability Statement

The data that support the findings of this study are not publicly available because the study included sensitive participant information but are available from the corresponding author upon reasonable request.

